# Sensorimotor deficits in distance runners with medial plantar pain

**DOI:** 10.1101/2021.02.16.21251637

**Authors:** Colton Funk, John J. Fraser, Jacob Resch, Jay Hertel

## Abstract

**Context:** Medial plantar pain is a complex and multifactorial condition experienced by some distance runners, which makes etiological differentiation and diagnosis challenging.

**Objective:** To assess plantar sensation, pain perception and sensitivity, intrinsic foot strength, and foot morphology before and after a 9.7 km run in long-distance runners with and without medial plantar pain.

**Design:** Descriptive laboratory study

**Setting:** Laboratory.

**Patients:** Seven distance runners with medial plantar pain (four males, three females; aged 22.3±3.7 years; BMI 22.3±3.5 kg/m^2^) and seven matched healthy controls (four males, three females; aged 20.3±1.0 years; BMI 22.0±1.7 kg/m^2^) were recruited from a public university.

**Intervention(s):** Participants ran a six-mile course in their own footwear at a self-selected pace.

**Main Outcome Measures:** Pain visual analogue scale (VAS), pressure pain thresholds (PPT), plantar sensation, foot morphology, weight-bearing dorsiflexion (WBDF), forefoot joint mobility, toe flexion, intrinsic foot muscle strength, and a seated neural provocation test were analyzed pre-and post-run utilizing mixed model group by time ANOVAs, *post hoc* effect size point estimates, and 95% confidence intervals.

**Results:** A significant group by time interaction was observed for PPT at the mid-arch (Control: pre: 83.0±27.4N, post: 79.5±22.6N; Symptomatic: pre: 90.5±31.9N, post: 70.1±32.7 N; p=.03) and posterior tibialis (PT) (Control: pre: 75.7±19.5 N, post: 65.7±14.2N; Symptomatic: pre: 75.8±20.4 N, post: 51.1±11.9 N; p=.05) sites. PPT in the posterior tibialis significantly decreased in the symptomatic group following the run (ES=1.5, 95% CI: 0.3, 2.7). Both groups demonstrated significantly decreased plantar sensation thresholds at the base of the 5^th^ metatarsal (p=.04), PPT at the calcaneal tuberosity (p=.001), and increased TMT extension (p=.01) and WBDF (p=.01) following the run.

**Conclusions:** The etiology of medial plantar pain observed in our sample was likely attributed to central sensitization and running-induced sensory changes. Clinicians should include sensorimotor testing when managing patients with medial plantar pain.

**Key Points:** Medial plantar pain is a unique pathologic entity that has not been previously well described and likely caused by central sensitization. Clinicians should include sensorimotor testing and interventions when managing patients with medial plantar pain.

## INTRODUCTION

Running-related lower extremity injuries are ubiquitous and affect about 20% of all runners, especially in individuals who run longer distances.^1^ With the increased popularity of long distance races such as half and full marathons, it is not uncommon for both recreational and elite runners to surpass 40 miles per week during training. The majority of running injuries occur at the foot and ankle, with plantar foot pain being a common complaint.^2^

There are several potential pain generators that may contribute to plantar foot pain in long distance runners. Factors such as foot posture and osseous malalignment of the tarsals that lead to intrinsic foot muscular demand may contribute to the onset and recovery of ankle-foot injury.^3^ Muscular fatigue during prolonged running may lead to altered running kinematics and kinetics^4^ and aberrant mechanical loading of osseous, ligamentous, muscular, and neurologic structures in the foot and ankle. As dynamic stabilizers of the medial longitudinal arch fatigue during cyclical loading, microtrauma to both dynamic and static stabilizers result in inflammation, abnormal collagen synthesis, and the onset of plantar pain.^5^ Repetitive mechanical stress may result in tissue failure when tension, compression, and shear exceeds the rate of physiological repair and remodeling.^6^

Due to the variability of injury mechanism and the multiple potential tissues affected, the clinical presentation of medial plantar pain in runners is complex. Plantar pain is the cardinal symptom in many conditions of the foot such as plantar fasciitis,^7^ tarsal tunnel syndrome,^8^ posterior tibialis tendinopathy,^9,10^ and stress fracture.^11^ These diagnoses share common mechanisms of injury as well as signs and symptoms that frequently present concurrently.However, plantar pain in the central portion of the medial longitudinal arch is a symptom distinct from the other running-related injuries and may have its own unique etiology.

The purpose of this descriptive study was to determine if there were differences in pain perception, pressure pain thresholds, plantar sensation, toe flexor and intrinsic foot strength, and foot morphology in long distance runners with and without medial plantar pain before and after a 9.7 km (6-mile) run.

## METHODS

### Design

The current study was a laboratory-based descriptive cross-sectional study. The independent variables were group (runners with medial plantar pain, asymptomatic controls) and time (pre-run, post-run) and the primary outcome measures were participant-reported pain and function, plantar sensation, hallux and lesser toe flexion strength, foot morphology across loading, and algometric pressure pain thresholds.

### Participants

Seven runners reporting primary plantar pain in the medial longitudinal arch (4 males, 3 females; aged 22.3±3.7 years; BMI 22.3±3.5 kg/m^2^) and 7 matched runners without medial plantar pain (4 males, 3 females; aged 20.3±1.0 years; BMI 22.0±1.7 kg/m^2^) participated in the current study (**Table 1**).

**Table 1.** Means and (Standard Deviations) for Participant Characteristics.

| Characteristics |  | Healthy<br>(n = 7) | Symptomatic<br>(n = 7) | (p-value) |
| --- | --- | --- | --- | --- |
| Demographics | Age, mean (SD), mo. | 20.3 (0.9) | 22.3 (3.7) | .21 |
|  | Height, mean (SD), cm | 166.6 (6.0) | 174.3 (11.1) | .14 |
|  | Weight, mean (SD), kg | 60.8 (4.6) | 67.1 (8.2) | .11 |
|  | BMI, mean (SD), kg*(m<sup>2</sup>)<sup>-1</sup> | 22.0 (1.7) | 22.3 (3.5) | .85 |
| Patient Reported Outcomes | # previous ankle sprains | 0.0 (0.0) | 0.7 (1.2) | .26 |
|  | # foot sprains | 0.0 (0.0) | 0.0 (0.0) | 1.00 |
|  | PROMIS Physical | 19.0 (0.6) | 17.4 (1.7) | .05 |
|  | PROMIS Mental | 17.4 (1.3) | 17.7 (1.8) | .74 |
|  | EQ5D | 0.8 (0.0) | 0.8 (0.1) | .34 |
|  | TSK 11 | 13.0 (2.8) | 16.0 (5.1) | .21 |
|  | Grit | 3.8 (0.6) | 4.2 (0.2) | .09 |
|  | FAAM Sport | 100.0 (0.0) | 89.4 (11.9) | .04 |
|  | FAAM SANE | 100.0 (0.0) | 95.0 (7.6) | .11 |
|  | FPI | 2.6 (2.4) | 0.4 (1.7) | .08 |
| Shoe Type | Trainers | 16.7% | 14.3% |  |
|  | Flats | 83.3% | 85.7% |  |
|  | Spikes | 0.0% | 0.0% |  |
| Foot Strike | Forefoot | 33.3% | 0.0% |  |
|  | Midfoot | 33.3% | 85.7% |  |
|  | Rearfoot | 33.3% | 14.3% |  |

\*Patient-Reported Outcomes Measurement Information System, European Quality of Life-5 Dimensions, Tampa Scale of Kinesiophobia-11, Short Grit Scale, Foot and Ankle Ability Measure-Sport, Foot and Ankle Ability Measure Single Assessment Numeric Evaluation, Visual Analog Score, Foot Posture Index-6

All participants met inclusion criteria as “long distance runners” if they ran at least 65 km (40 miles) per week. Runners were included in the symptomatic group if they experienced a primary complaint of medial plantar foot pain that was exacerbated from running. Participants with comorbid plantar fasciosis,^7^ or posterior tibialis tendinitis^9,10^ were included only if these symptoms were secondary to the primary focal medial plantar pain. Participants in the asymptomatic control group were matched by sex and test limb and were included if they had no complaint of plantar foot symptoms while running. Individuals were excluded if they had signs of tarsal tunnel syndrome,^8^ osseous,^12–14^ or ligamentous injury,^12^ or reported a history ankle or foot sprain in the past year, fracture or surgery in the leg or foot, self-reported disability resulting from lower extremity pathology, neurological or vestibular disorders that affected balance, diabetes mellitus, lumbosacral radiculopathy, a soft tissue disorder such as Marfan or Ehlers-Danlos syndrome. Participants provided informed consent and the study was approved by the university’s Institutional Review Board for Health Science Research.

### Screening Procedures

Following consent, participants provided medical history and completed the PROMIS General Health Questionnaire version 1.1,^15^ Foot and Ankle Ability Measure-Sport Scale,^16^ the Tampa Scale of Kinesiophobia-11 item scale,^17^ and the Grit Scale of Resilience.^18^ Participants were asked to rate the severity of their foot pain at the present and at the worst in the past week using a 10-centimeter visual analogue scale (VAS). If participants experienced symptoms in both feet, the foot that was more problematic was evaluated. A board certified orthopedic physical therapist with 15-years of clinical experience completed a screening consisting of observation for the posterior tibial edema sign,^10^ plantar arch ecchymosis,^12^ or the gap sign;^12^ a palpatory examination of the proximal plantar fascia,^7^ tarsal tunnel,^8^ posterior tibial tendon,^10^ the Ottawa Ankle Rule screening;^14^ the single limb heel raise test;^9^ and performance of provocative tests that include the windlass test,^7^ Tinel’s test of the tarsal tunnel,^8^ and the single leg hop test.^13^ Following screening, an athletic trainer with one year clinical experience, who was blinded to clinical history conducted pre-and post-run assessments.

### Neurosensory Assessment Procedures

Sensory testing was performed at the plantar aspect of the heel, the base of the 5^th^ metatarsal, and the head of the 1^st^ metatarsal using Semmes-Weinstein Monofilaments (Smith & Nephew, Inc, Germantown, WI) in the 4-2-1 stepping protocol.^19^ Pressure pain thresholds were tested with a push-pull force gauge (FDN 100, Wagner Instruments, Greenwich, CT) at the apex of medial longitudinal arch, plantar fascia origin, and posterior superior medial malleolus.^20^ Participants were instructed to verbalize when the applied pressure reached an intolerable level. Each location was tested three times with at least 30 seconds between trials with the average of the three trials recorded. Passive seated knee extension was tested with the foot dorsiflexed to assess neural provocation and plantarflexed to assess hamstring length^21^ using a digital inclinometer (Fabrication Enterprises, White Plains, NY). The participant was seated at the foot of the examination table, while sitting erect and holding a dowel behind their back with the occiput, thoracic spine, and the sacrum in contact. The average of three trials was recorded.

### Ankle-Foot Morphology, Mobility, and Neuromotor Assessment Procedures

Foot posture was assessed using the Foot Posture Index-6 (FPI-6).^22^ Total foot length, truncated foot length, foot width, and dorsal arch height were measured seated, double limb standing, and single limb standing using the Arch Height Index Tool (JAKTOOL, Inc, Cranberry, NJ).^22^ Arch flexibility and foot mobility magnitude were calculated from the morphologic foot measurements across loading. Ankle dorsiflexion was assessed using the weight bearing dorsiflexion test.^22^ The intrinsic foot muscle (IFM) test was performed as described by Jam.^23^ Forefoot inversion and eversion motion was measured using a digital inclinometer (Fabrication Enterprises, White Plains, NY) as described by Fraser and colleagues.^22^ First metatarsal dorsiflexion and plantar flexion were measured using a custom device consisting of two bent rulers.^22^ Forefoot and 1^st^ tarsometatarsal measures were performed three times, with the average of each motion recorded. Three trials of hallux and lesser toe flexor strength were assessed using a microFET2 digital handheld dynamometer (Hoggan Health Industries, West Jordan, UT),^22^ with the highest force recorded.

### Intervention

Following the baseline assessment, participants ran an outdoor 9.7 km running course in their own footwear. Participants were instructed to run at a pace that was habitual and based on their current level of training. Immediately following the run, each participant was asked to rate their current foot pain using the VAS. A post-run assessment consisting of sensory testing, foot morphology, intrinsic foot muscle test, toe flexion strength, pressure pain threshold, seated knee extension, forefoot inversion and eversion, 1^st^ metatarsal dorsiflexion and plantar flexion, and weight-bearing dorsiflexion range of motion tests was performed. Participants were dismissed from the study following their post-run assessment.

### Statistical Analysis

Descriptive statistics were calculated for group demographic and self-reported measures, with independent t-tests used to assess group differences. Proportion estimates and 95% CI were calculated for all dichotomous variables. The Wilcoxon’s signed rank test was used to assess group differences for the IFM test at pre-and post-run time points. Ordinal measures that had greater than five items (plantar sensation) were treated as continuous data during analysis.^24^ Mixed model group by time ANOVAs and *post hoc* Cohen’s *d* effect size (ES) point estimates and 95% confidence intervals (CI) were calculated. ES were interpreted using the scheme proposed by Cohen:^25^ as trivial (<0.2), small (0.2-0.49), moderate (0.5-0.79), or large (>0.8). Pre-to-post treatment ES point estimates and 95% CI were statistically significant when the CI did not cross the ‘0’ threshold. Data was analyzed using Statistical Package for Social Sciences (SPSS) Version 23.0 (IBM, Inc., Armonk, New York). Proportion point estimates, Cohen’s *d* effect sizes, and 95% confidence intervals were calculated using Microsoft Excel for Mac 2016 (Microsoft Corp., Redmond, WA). The level of significance was p ≤ 0.05 for all analyses.

## RESULTS

No statistical differences were observed for demographics, injury history, or foot posture between groups. The symptomatic group had significantly decreased PROMIS Physical composite and FAAM Sport scores compared to controls (**Table 1)**. Both groups primarily utilized training shoes during long distance running. Running style was equally distributed among the control group (rearfoot: 33.3%, midfoot: 33.3%, forefoot: 33.3%). The majority (85.7%) of the symptomatic group was observed to have a midfoot strike pattern.

A significant group by time interaction for pressure pain threshold at the mid-arch (Control: pre: 83.0±27.4 N, post: 79.5±22.6 N; Symptomatic: pre: 90.5±31.9 N, post: 70.1±32.7 N; p=.03) and posterior tibialis (Control: pre: 75.7±19.5 N, post: 65.7±14.2 N; Symptomatic: pre: 75.8±20.4 N, post: 51.1±11.9 N; p=.05) sites was present (**Table 2**). Only the posterior tibialis in the symptomatic group demonstrated a significantly large decrease (ES=1.5, 95% CI: 0.3, 2.7) in pressure pain threshold following the run (**Figure 4**). Both groups demonstrated decreased thresholds for plantar sensation at the base of the 5^th^ metatarsal (p=.04) and pressure pain at the calcaneal tuberosity (p=.001) following the run (**Table 2**) and a significant increase in TMT extension (p=.01) and WBDF (p=.01) (**Table 3**). There were no significant differences in IFM test performance pre-to-post run in either group (Control: pre: 2.4±0.8, post: 2.7±0.5, p=.16; Symptomatic: pre: 2.4±0.8, post: 2.4±0.8, p=1.00). There were no additional significant group or time effects or group by time interactions.

**Table 2.** Means and (Standard Deviations) for Neurosensory Assessment.

|  | Healthy |  | Symptomatic |  | Group | Time | Group x Time |
| --- | --- | --- | --- | --- | --- | --- | --- |
| Dependent Variable | Pre | Post | Pre | Post | (p-value) | (p-value) | (p-value) |
| **PPT Mid-Arch, mean (SD), N** | 83.0<br>(27.4) | 79.5<br>(22.6) | 90.5<br>(31.9) | 70.1<br>(32.7) | .95 | .005 | .03 |
| **PPT Calcaneal Tuberosity, mean (SD), N** | 110.1<br>(26.7) | 100.6<br>(21.5) | 102.4<br>(31.6) | 85.6<br>(34.5) | .47 | .001 | .25 |
| **PPT Posterior Tibialis, mean (SD), N** | 75.7<br>(19.5) | 65.7<br>(14.2) | 75.8<br>(20.4) | 51.1<br>(11.9) | .41 | <.001 | .05 |
| **Monofilament Heel, mean (SD), g** | 15.7<br>(26.5) | 8.7<br>(10.0) | 9.5<br>(5.8) | 10.1<br>(9.4) | .74 | .41 | .33 |
| **Monofilament Base of 5<sup>th</sup>, mean (SD), g** | 3.1<br>(3.9) | 1.8<br>(1.9) | 4.7<br>(3.9) | 1.8<br>(1.8) | .55 | .04 | .42 |
| **Monofilament 1st metatarsal head, mean (SD), g** | 8.5<br>(10.4) | 5.5<br>(6.6) | 8.2<br>(5.9) | 5.6<br>(5.4) | .97 | .22 | .93 |

SD=standard deviation, N=Newtons, g=grams

**Table 3.** Means and (Standard Deviations) for Ankle-Foot Morphology, Mobility, and Neuromotor Assessment.

| Dependent Variable | Healthy |  | Symptomatic |  | Group<br>(p-value) | Time<br>(p-value) | Group x Time<br>(p-value) |
| --- | --- | --- | --- | --- | --- | --- | --- |
|  | Pre | Post | Pre | Post |  |  |  |
| Foot mobility magnitude,<br>mean (SD), cm | 2.5<br>(1.0) | 1.7<br>(0.9) | 2.2<br>(1.5) | 2.1<br>(1.6) | .92 | .08 | .21 |
| Arch Flexibility, mean (SD),<br>cm*kg<sup>-1</sup> | 0.04<br>(0.01) | 0.03<br>(0.01) | 0.03<br>(0.01) | 0.03<br>(0.01) | .08 | .20 | .43 |
| TMT extension, mean (SD),<br>mm | 1.5<br>(1.0) | 2.3<br>(1.0) | 2.3<br>(1.9) | 2.6<br>(1.8) | .51 | .01 | .16 |
| TMT flexion, mean (SD), mm | 4.1<br>(1.8) | 3.9<br>(1.9) | 4.2<br>(1.6) | 4.1<br>(0.7) | .81 | .62 | 1.00 |
| Forefoot inversion, mean<br>(SD), degrees | 33.2<br>(7.7) | 31.4<br>(5.0) | 32.4<br>(2.2) | 32.8<br>(3.5) | .90 | .53 | .33 |
| Forefoot eversion, mean<br>(SD), degrees | 14.0<br>(2.6) | 13.5<br>(4.2) | 16.3<br>(3.6) | 15.3<br>(2.1) | .18 | .44 | .75 |
| WBDF, mean (SD), cm | 10.4<br>(4.2) | 11.8<br>(3.5) | 11.7<br>(2.2) | 12.2<br>(2.4) | .62 | .01 | .12 |
| Lesser toe flexion, mean<br>(SD), Nm*kg<sup>-1</sup> | 128.8<br>(25.6) | 124.6<br>(11.3) | 120.0<br>(31.4) | 122.0<br>(37.7) | .69 | .86 | .62 |
| Hallux flexion, mean (SD),<br>Nm*kg<sup>-1</sup> | 141.4<br>(28.1) | 136.5<br>(24.6) | 142.0<br>(54.5) | 131.1<br>(46.6) | .91 | .28 | .68 |
| Knee Ext. with DF, mean<br>(SD), degrees | 22.2<br>(10.6) | 20.7<br>(11.1) | 18.1<br>(7.8) | 19.9<br>(8.1) | .63 | .90 | .14 |
| Knee Ext. with PF, mean<br>(SD), degrees | 13.8<br>(9.0) | 12.7<br>(9.6) | 10.5<br>(5.5) | 11.5<br>(6.1) | .59 | .17 | .95 |

SD=standard deviation, cm=centimeters, cm\*kg<sup>-1</sup>=centimeters per kilogram, mm=millimeters, Nm\*kg<sup>-1</sup>=Newton-meters per kilogram

**Figure 1.**
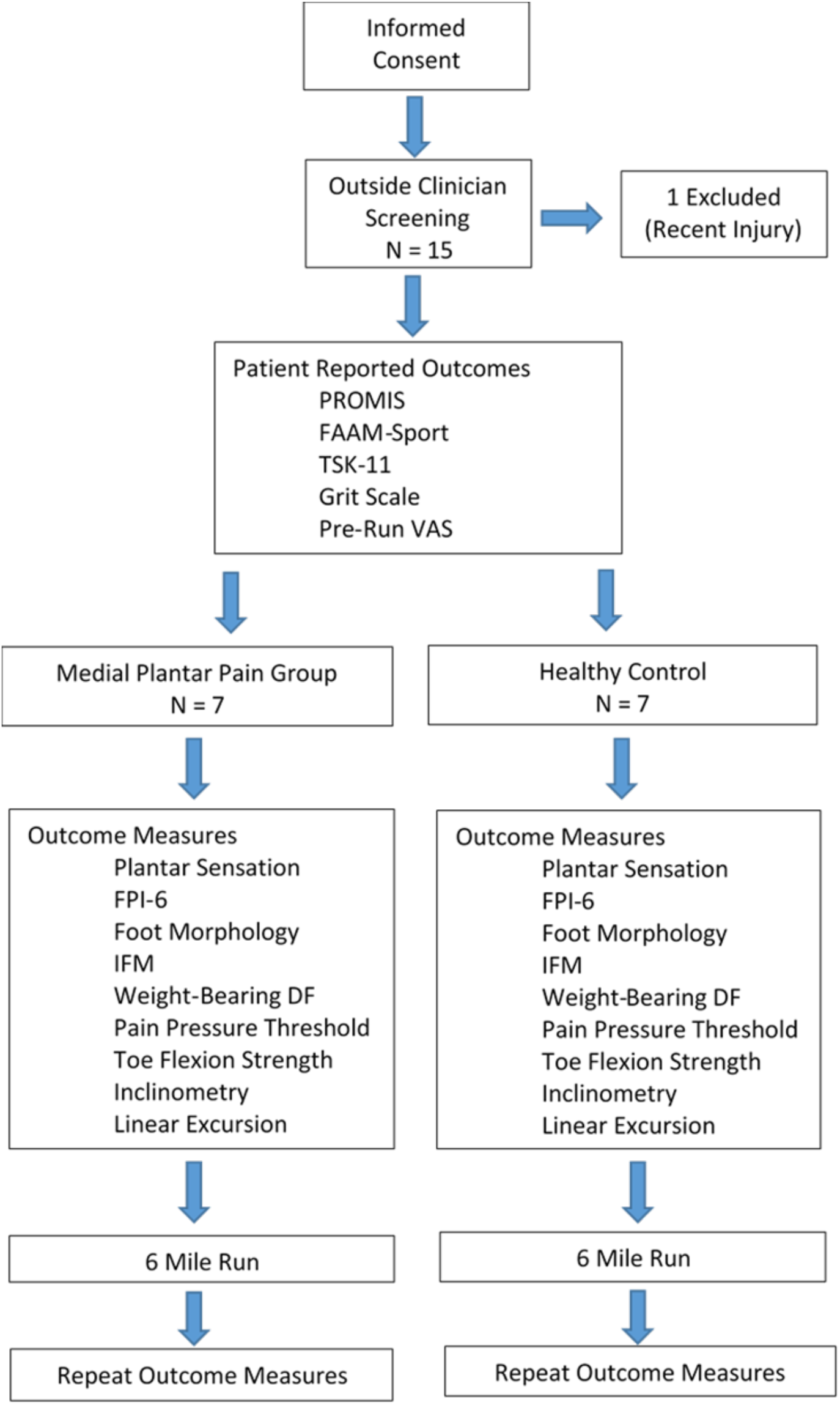
CONSORT Flow Chart.

**Figure 2.**
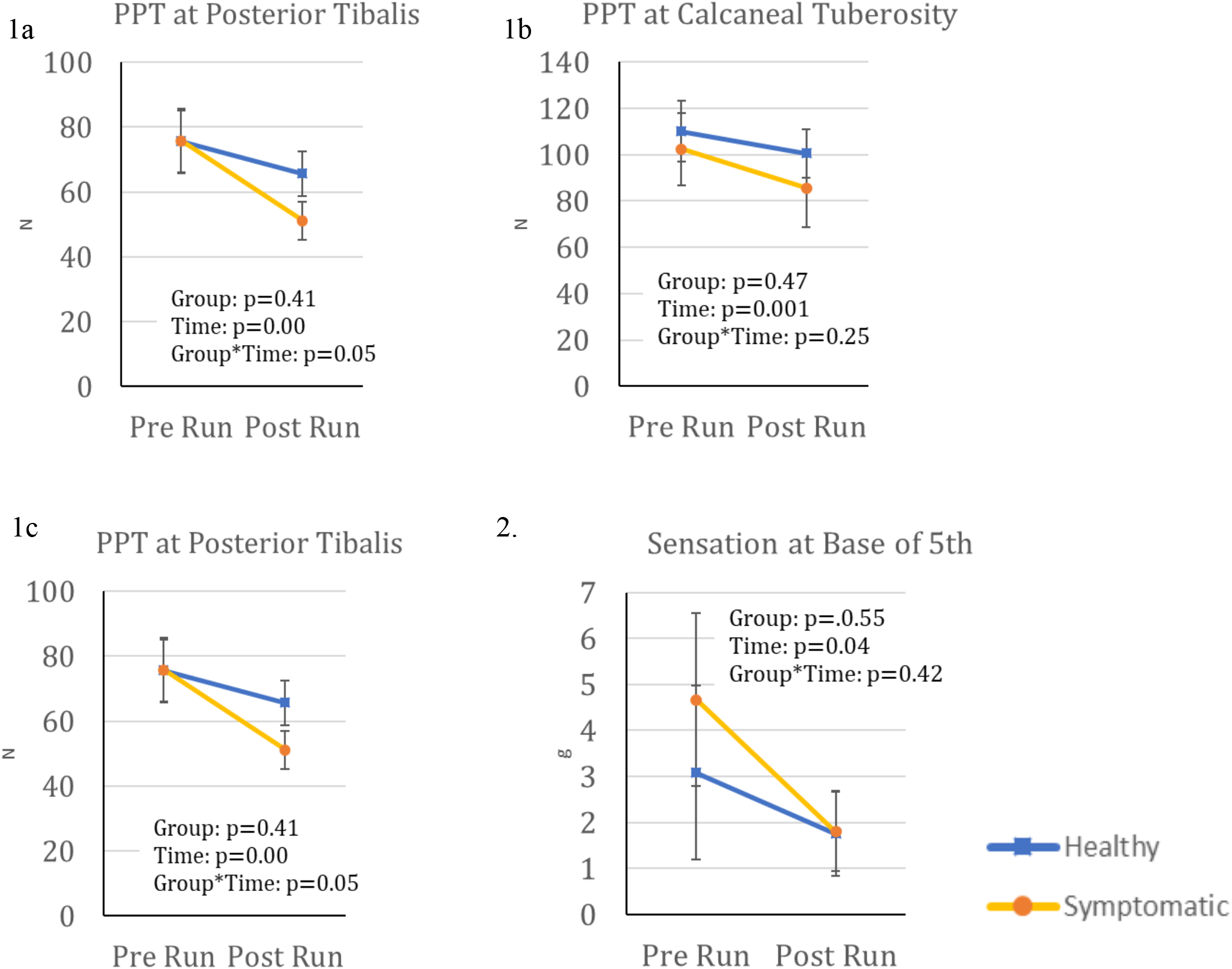
Pre and post measures of 1. pressure pain thresholds (PPT), measured in Newtons, at the three locations: apex of the medial longitudinal arch, b) calcaneal tuberosity, c) posterior to the medial malleolus along the posterior tibialis tendon and 2. plantar sensation, measured in grams, at the base of the 5^th^ metatarsal

**Figure 3.**
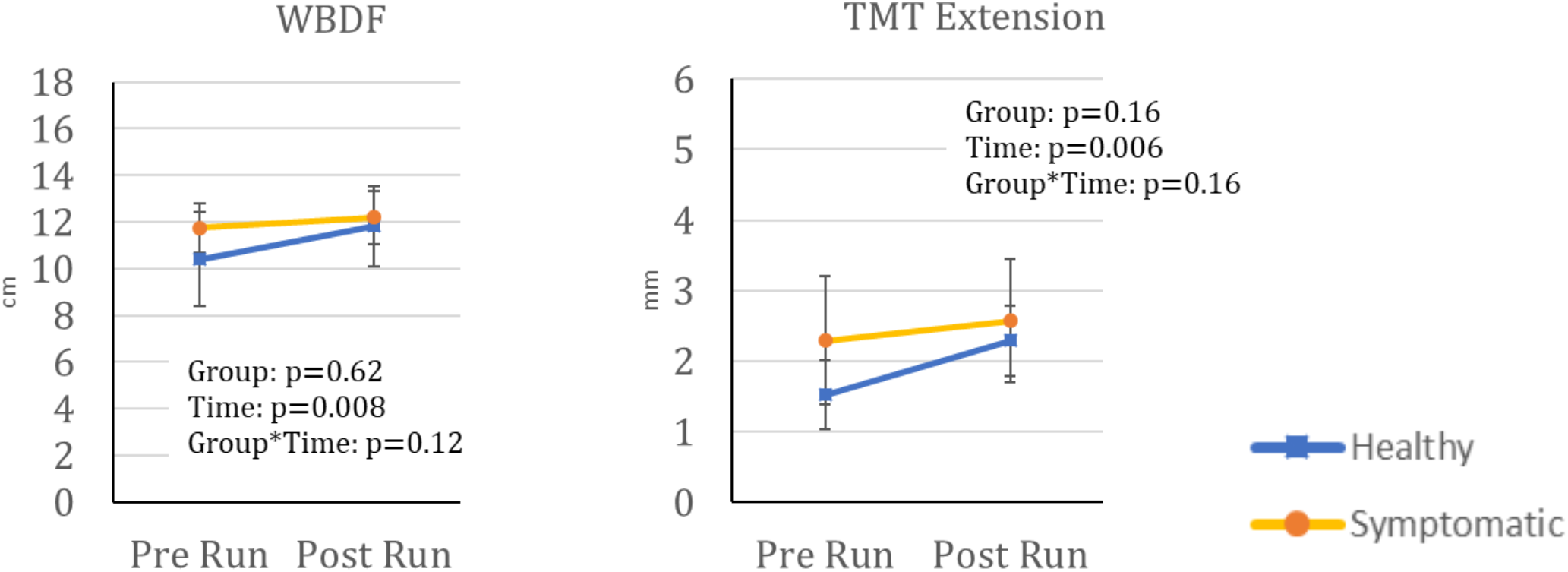
Pre and post measures of weight bearing dorsiflexion (WBDF) and tarsometatarsal (TMT) extension range of motion measures.

**Figure 4.**
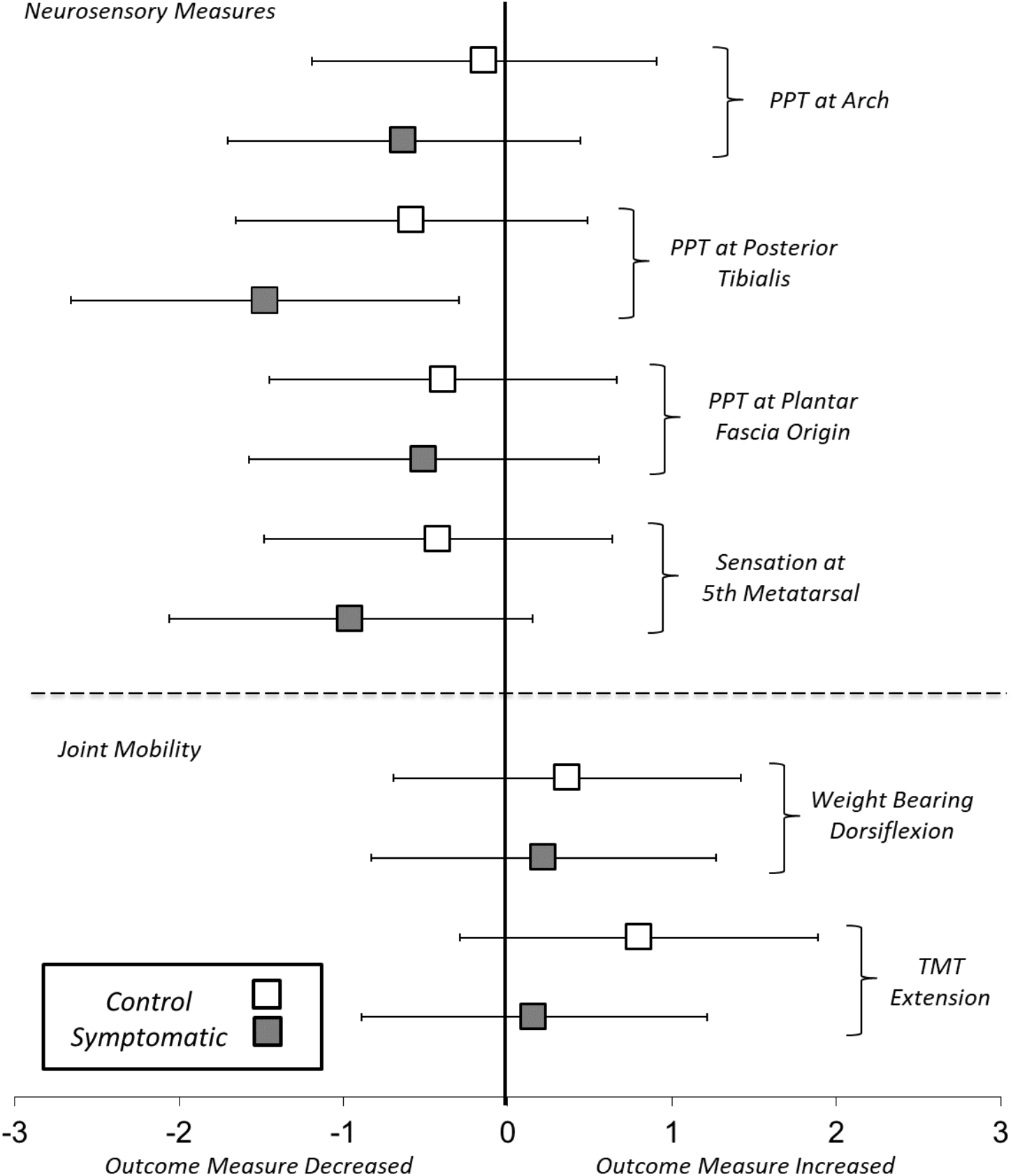
Pre-to-post run effect size estimates (Cohen’s d) and 95% confidence intervals.

## DISCUSSION

To the author’s knowledge, the current study is the first to describe neurophysiologic changes in long distance runners with a primary complaint of medial plantar pain. The primary findings of this case control study were that runners with medial plantar pain had a significant decrease in pressure pain threshold prior to and following a 9.7 km (6-mile) run at the apex of the arch and the posterior tibialis muscle compared to a similarly matched control group that did not experience a change. Both groups demonstrated a significantly decreased threshold in pressure pain at the plantar fascia origin and sensation at the base of the 5^th^ metatarsal and increased 1^st^ tarsometatarsal dorsiflexion, and weight bearing dorsiflexion motion following their run.

### Neurosensory Function

Decreased pressure pain thresholds in the apex of the arch and the posterior tibialis muscle after the run is likely attributed to a combination of local nociception of tissue reactivity originating in the medial longitudinal arch and central neurosensory effects. The decreased pressure pain threshold at the posterior tibialis had a large effect size (**Figure 4**), supporting central sensitization as a plausible explanation.

Central sensitization is a phenomenon that results from up regulation of afferent nociceptive pathways in the spinal dorsal horn which is facilitated by pain generation in adjacent pain pathways.^26^ This nociceptive up regulation results in regional hyperalgesia and remote sensory changes away from the primary pain generator.^26^ The medial longitudinal arch is innervated by the medial plantar nerve, a branch of the tibial nerve. Since the posterior tibialis shares the same innervation proximally in the deep posterior compartment of the leg, it is plausible that the observed proximal changes in pressure pain thresholds were the results of central sensitization caused by pain generators in the medial plantar foot.

It is also plausible that the symptomatic group had a peripheral neuropathic contributor to their symptoms. While conceivable, our findings do not support this mechanism. Participants with medial plantar pain did not present with paresthesia, sensory loss, and or motor loss during the heel raise screening or strength testing, alterations of foot morphology across loading, or tibial nerve provocation before or following the 9.7 km run.

Healthy and symptomatic groups experienced decreased pressure pain thresholds in the plantar fascia and decreased sensory thresholds at the plantar aspect of the 5^th^ metatarsal base following the run. These findings are likely related to physiological sensory changes that have been associated with exercise. Hosseinzadeh et al.^27^ found pressure pain thresholds decrease after a repeated bout of eccentric exercise, suggesting a hypersensitization of local soft tissue. It is also conceivable that variability of sensory testing with monofilaments or anticipation of pain during algometric assessment may also explain our findings.

### Ankle-Foot Morphology, Mobility, and Neuromotor Strength

We observed an absence of group differences or changes in foot morphology, midfoot joint motion, or strength after the run. These results are suggestive that medial arch pain in our sample was not a mechanical problem. Previous research has theorized that decreased medial longitudinal arch height may induce a greater strain on the supporting structures of the foot that may result injury.^5^ Our findings do not substantiate this supposition.

The observed increase in ankle dorsiflexion following the our participants’ run is consistent with changes found following dynamic warm-up exercise.^28^ Exercise has previously been demonstrated to increase muscle and connective tissue temperature and therefore increase tissue pliability.^28^ While we did not assess muscle temperature in our study, we can assume that our participants experienced increased core temperature following the exercise that likely resulted in increased tissue elasticity.

### Clinical Implications

Our results identified clinical characteristics of medial plantar pain in long distance runners. Clinicians should be cognizant that medial plantar pain is complex, may encompass multiple potential pain generators, and may involve central pain mechanisms. We recommend that clinicians take a holistic approach during examination of these patients and assess the multiple segments of the ankle-foot complex. Neurosensory testing that includes monofilament testing and pressure pain thresholds may have clinical utility in the assessment of these patients and should be performed as part of a comprehensive evaluation in patients presenting with medial plantar pain.

We recommend that clinicians assess running gait mechanics in patients with medial plantar pain. Daoud and colleagues^29^ found runners with rearfoot strike patterns to have higher rates of injury compared to forefoot strike patterns. However, in our study, we found that 85.7% of the symptomatic group were midfoot strikers. Therefore, it is plausible that midfoot strike patterns may play a role in the pathomechanics that lead to medial plantar pain. These results should be interpreted with caution because of our small sample size and should be confirmed in a larger, more diverse sample of distance runners.

While it is unclear if decreased sensory thresholds are protective or contributory to pain symptoms, we speculate that it may be beneficial for patients with medial plantar pain to perform exercises barefoot on a variety of different textured surfaces such as carpet, foam, turf, grass, or sand. Repeat exposure to different surfaces may desensitize these patients through habituation and sensory reeducation. Manual therapy, to include myofascial release, soft tissue mobilization, and joint mobilization, should be considered as a treatment modality in the management of medial plantar pain due to central neurophysiologic effects in reducing pressure pain thresholds^20^ and cutaneous hypersensitivity^30^ in the foot.

### Limitations and Future Considerations

The study is not without limitations. It is unclear whether sensory changes preceded the development of medial plantar pain or if this was a consequence following the development of symptoms. Our preliminary study consisted of a small convenience sample. Combined with high variability of some of the measures, it is plausible that we were underpowered to establish statistically significant differences for some of the mechanical outcome measures. Future research using a larger sample is needed. We also did not control for orthotic use, running shoe type, or time to complete the run. While these delimitations may be viewed as a potential limitation, they improve generalizability. Future studies of the effects of sensitization and pressure pain changes at increased distance, such as half marathon or marathon distance is recommended.

### Conclusion

Medial plantar pain is a unique pathologic entity that has not been previously well described and likely caused by central sensitization. We found that long distance runners with medial arch pain had a greater change in pressure pain thresholds at the arch and at the distal posterior tibialis muscle compared to healthy participants after a 6-mile run. There were no changes in toe flexor strength, foot motion, or tibial nerve provocation after the run suggesting central sensitization plays a role in this clinical entity. Clinicians should include sensorimotor testing and interventions when managing patients with medial plantar pain.

## Data Availability

Due to the nature of this research, participants of this study did not agree for their data to be shared publicly, so supporting data is not available.

